# Enhancement of *M. tuberculosis* Line Probe Assay Sensitivity through Whole Genome Amplification of Low-Quantity DNA Released from Sputum and Archived on Chemically-Coated Cellulose Matrix Using an Isothermal Enzymatic Strand-Displacement Process

**DOI:** 10.1101/2024.06.27.24309624

**Authors:** Krishna H. Goyani, Chirajyoti Deb, Daisy Patel, Shalin Vaniawala, Pratap N. Mukhopadhyaya

## Abstract

In this study, thirty-nine sputum samples from tuberculosis (TB)-positive patients undergoing first-line therapy were collected and archived on a chemical-coated cellulose matrix. DNA was extracted from these matrices and tested for *Mycobacterium tuberculosis* using the Xpert MTB/RIF Ultra assay. Seven samples tested positive for *M. tuberculosis*, with low levels of detection. End-point PCR yielded faint signals in four samples, but no signal in the remaining three. A Line Probe Assay (LPA) detected pathogen DNA in only one of the three PCR-negative samples. Remarkably, the LPA-negative samples were successfully detected by LPA and end-point PCR following isothermal, strand displacement-based whole genome amplification (WGA) of the stock DNA. The drug sensitivity profile of these samples was consistent with the Xpert MTB/RIF Ultra results obtained from the original stock DNA. Additionally, sputum DNA from healthy individuals spiked with 125 ng, but not with 1.25 ng, of *M. tuberculosis* culture DNA was detected by LPA. Following WGA, the 1.25 ng sample was also detected successfully by LPA, and its drug sensitivity profile matched that of the 125-ng sample. These findings indicate that WGA of sputum DNA from a cellulose matrix, even with low pathogen load, can enhance the detection capabilities of LPA by enriching the genome target through an isothermal enzymatic strand displacement method. This method holds promise for improving diagnostic sensitivity in TB detection.

## Introduction

Tuberculosis is a dreaded infectious disease with worrisome statistics associated with it. It causes around 10.4 million cases and 1.8 million events of mortality in a year across the globe. It is estimated that around 4.3 million infected persons fail to get diagnosed [1]. The complexity of this disease compounds due to rapid emergence of multidrug and extensively drug-resistant tuberculosis (MDR-TB and XDR-TB, respectively) that poses a major challenge toward efforts for worldwide control of tuberculosis [2].

Culture based methods of testing drug susceptibility of M. tuberculosis using solid media is an effective and reliable technique. But one of the major shortfalls of this method is its long turnaround time to produce results which may be between 8 and 12 weeks [3]. A relatively faster and sophisticated alternative is the liquid-based culture methods which too may take a long turnaround time of 4 to 6 weeks [4]. This long period of time to generate results is detrimental since it translates to long periods of nil or ineffective therapy which promote rampant transmission of the pathogen. This is the reason for rapid evolution of molecular techniques for detection of *M. tuberculosis* and its drug-resistant variants [5]. The turnaround time here is often less than a day and the methods generate reliable results.

In recent times, line probe assays or LPAs have emerged as a reliable and rapid molecular method of detection of *M. tuberculosis*, particularly their drug resistant variants. Despite the complexity associated with the protocol, they reliably detect isoniazid (INH) as well as rifampicin (RIF) resistance in *M. tuberculosis* and are comparatively less expensive compared to Xpert MTB assays (Cepheid, Sunnyvale, CA, USA). Further, this test concludes in an average mean time of around 8 hours [6].

In line probe assay, the DNA from a clinical sample is extracted using conventional but rapid methods and the species-specific as well as drug resistant determining regions within the *M. tuberculosis* genome are amplified with oligonucleotide primers tagged with biotin (Biotinylated probes). These labelled PCR amplicons are then hybridized to specific probes and immobilized on a solid support in the form of thin strips. These trapped hybrids are then detected using the classical biotin – avidin alkaline phosphatase detection protocol [7] by colorimetry. The final result is in the form of thin coloured lines that are scorable by naked eyes.

The line probe assay from Hain Life science has clinically useful variants. The GenoType MTBDRplus kit detects *M. tuberculosis* complex as well as its resistance to rifampicin and/or isoniazid from clinical specimens and cultures while GenoType MTBDRsl kit is for detection of resistance to fluoroquinolones, aminoglycosides/cyclic peptides and/or ethambutol from *M. tuberculosis* clinical specimens and cultures (Hain Lifescience GmbH, Nehren, Germany).

However, one of the shortfalls of line probe assay is its relatively low level of sensitivity when compared to other molecular assays in its league such as Xpert MTB Rif Ultra (Sunnyvale, CA, USA). Studies indicate that the limit of detection of the LPA is around 10,000 CFU/ml [8] which is similar to smear microscopy. In comparison, culture-based method of detection of drug sensitivity in *M tuberculosis* is a low of 10-100 CFU/ml [9]. It is likely that LPA is prone to missing samples with a low bacillary load. This is particularly visible when patients are on first-line antituberculosis treatment during the time of collection of samples that often affects the quantity of target DNA in clinical samples. [10]

In recent years, techniques and approaches have evolved for superior methods of collection, transportation, storage and release of DNA from *M. tuberculosis*-infected clinical samples with special reference to sputum. Goyani et al (2023) described a chemically coated cellulose matrix that could be effectively used for sputum storage and DNA-release devices and with the advantage of optimal biocontainment properties [11]. The device demonstrated compatibility with cartridge-based nucleic acid amplification test platforms such as Xpert MTB Rif Ultra and/or its variants (Sunnyvale, CA, USA) [12] which are known for their high level of sensitivity and specificity [13]. However, when reagent-coated matrixes such as the one cited above are used for archival and release of DNA from sputum containing low quantum of infected *M. tuberculosis* bacilli the low quantity of DNA result in false negative results in line probe assay. In this study, we demonstrate use of whole genome amplification of the heterogeneous DNA population released from human sputum DNA archived in a reagent coated cellulose matrix [11] to convert false negative calls for low target DNA-containing samples to true positive ones by artificially increasing the quantum of available DNA for line probe assay to detect *M. tuberculosis* drug resistance [14].

## Materials and Methods

### Clinical samples of M. tuberculosis

A set of 39 clinical sputum samples from patients aged 25-40 years, diagnosed with tuberculosis and undergoing first-line therapy, along with 3 clinical samples from healthy individuals, were obtained from Microcare Laboratory and Tuberculosis Research Centre in Gujarat, India, between January and December 2023. The sputum samples were spotted onto a reagent-coated cellulose matrix, followed by DNA release and screening using the Xpert MTB/Rif assay (Sunnyvale, CA, USA) as described by Goyani et al. (2023) [11]. A panel of seven *M. tuberculosis*-positive samples and three negative samples was created. The positive samples, coded as XLw-1 through XLw-7, were classified as ‘MTB detected, low,’ according to the classification provided by the Xpert MTB Rif Ultra assay, indicating a low level of *M. tuberculosis* DNA in the reaction. The negative samples, coded as D1 through D3, were classified as ‘MTB not detected.’

### Ethical considerations

This study was conducted in accordance with the ethical standards outlined in the Declaration of Helsinki and received approval from the Institutional Review Board of Nirmal Hospital (Approval Number: Nirmal/HPL/Ethics/001). Written informed consent was obtained from all participants prior to their inclusion. The procedures were designed to ensure the confidentiality and anonymity of the participants throughout the research process. Additionally, the study received consent from the Department of Biotechnology, Government of India-recognized Institutional Biosafety Committee (IBSC) of Wobble Base Bioresearch Private Limited, with approval number WBB/IBSC/TBSC-03.

### Release of DNA for Line Probe Assay (LPA)

The reagent-coated cellulose matrix containing clinical samples identified as ‘Low’ positive by the Xpert MTB Rif assay underwent a 48-hour incubation at room temperature to release DNA. The membrane was soaked in sterile double-distilled water for 15 minutes. Subsequently, 650 μL of the resulting solution was transferred to a sterile 2 ml centrifuge tube and combined with 65 μL of 3 Molar Sodium Acetate (Sigma-Aldrich, St. Louis, MO, USA). DNA precipitation was achieved by adding 1.4 ml of ice-cold 100% Ethyl alcohol, followed by centrifugation at 4°C for 5 minutes. The DNA pellet was washed with 70% Ethyl alcohol, air-dried, and resuspended in 50 μL of sterile nuclease-free water. Five μL of this DNA solution was utilized for both line probe and endpoint PCR assays.

### Processing of DNA for Line Probe Assay (LPA)

The Line Probe Assay was performed according to the manufacturer’s instructions (Hain Lifescience GmbH, Nehren, Germany). Initially, all reagents were thawed on ice, and the PCR reaction mixture was prepared as per the kit’s protocol. Subsequently, 5 μL of precipitated DNA was added to the PCR reaction mixture, which was then run in a designated thermal cycler program.

For hybridization and detection, PCR products were denatured by heating and promptly transferred to an ice-cold hybridization buffer, where they were applied to test strips in a hybridization tray. Following this step, incubation in a water bath at the specified temperature was conducted, followed by a rigorous wash to remove nonspecific DNA molecules.

Processed strips were then incubated with a conjugate reagent, underwent a washing step, and were re-incubated in substrate solution for result visualization. A final wash was performed to stabilize the signals. Finally, the strips were dried, and the results were interpreted by comparing them with the reference guide provided with the kit.

### End Point PCR amplification of precipitated DNA

For end point PCR, 5 μL of precipitated DNA was used. Ten picomoles of forward and reverse primers [CP089781.1: g. 1470368 to g.1470387 (+) and CP089781.1: g.1470567 to g.1470588 (-) respectively] were added to a commercial Thermo Scientific PCR Master Mix (Thermo Fisher Scientific, USA) diluted to 1 X concentration with nuclease free water. The target DNA was amplified in a total volume of 25 μL on a Perkin Elmer / Applied Biosystems GeneAmp PCR System 9600. The thermal cycling conditions were as follows: 94^0^C – 2 minutes (hold), 94^0^C - 30 seconds, 58^0^C - 30 seconds and 72^0^C - 30 seconds (35 cycles), 72^0^C – 2 minutes (hold) and 4^0^C – 5 minutes (hold). Five μL of the PCR products were resolved on a 2% agarose gel (Seakem agarose (Lonza, Basel, Switzerland), stained with Ethidium bromide (Sigma-Aldrich, St. Louis, MO, USA) at a final concentration of 0.5 μg/ml and the bands recorded using a Canon EOS Rebel T7i digital camera with an orange filter. The gel images were analysed using the GelAnalyzer 23.1.1 software (www.gelanalyzer.com).

### Formulating dilutions of M. tuberculosis DNA-spiked clinical samples

M. tuberculosis DNA was extracted from cultures grown in Middlebrook 7H10 media [15] using the QIAamp DNA Mini Kit (Qiagen, Hilden, Germany). Sputum DNA from samples D1, D2, and D3 (from healthy individuals) were combined in equal proportions. The extracted *M. tuberculosis* DNA was added to 1 ml of *M. tuberculosis*-negative sputum DNA mix, and two different dilutions of the extracted DNA were prepared.

In Dilution-1 stock DNA, the concentration of *M. tuberculosis* DNA was 25 ng/μL, while in Dilution-2 stock DNA, it was 0.25 ng/μL, respectively. Five μL of these dilutions, totalling 125 ng and 1.25 ng, respectively, were used to set up LPA or whole genome amplification reactions. All experiments were performed in triplicate to confirm the repeatability of the results, unless otherwise specified.

### Whole Genome Amplification (WGA) of the cellulose matrix extracted sputum DNA and M. tuberculosis DNA-spiked clinical samples

For whole genome amplification of the heterogeneous DNA released from the reagent coated cellulose matrix spotted with *M. tuberculosis* positive sputum or the sample (Dilution-2) formulated by spiking *M. tuberculosis* culture DNA to *M. tuberculosis*-negative sputum DNA, 5 μL of the DNA was used in a 25 μL reaction volume. The reaction composed of Tris-HCl (pH 8.8): 40 mM; KCl: 20 mM; (NH_4_)_2_SO_4_: 20 mM; MgSO_4_: 6 mM; dNTPs: 1.4 mM each; *Bst* DNA Polymerase (New England Biolabs (NEB): 16 units; Random primer: 5 μM (Thermo Fisher Scientific (Catalogue No. 48190011) and Betaine: 1 M final concentration. The incubation was performed on Perkin Elmer / Applied Biosystems GeneAmp PCR System 9600 and the incubation temperature was as follows: 95^0^C-30 seconds and 65^0^C for 10 minutes.

## Results

DNA from sputum samples, archived in the reagent-coated cellulose matrix and corresponding to 7 low-positive Xpert MTB Rif Ultra reports were extracted and subjected to end point PCR (25 μL reaction volume) using ITS (Internal Transcribed Spacer) region of the *M. tuberculosis* genome as the target. Out of the 7 samples, 4 generated faint bands (Samples XLw-4 - XLw-7) of desired molecular weight while the remaining 3, labelled as XLw-1 - XLw-3 did not generate any bands (Figure 1). Inhibition-check PCR using an external DNA and its corresponding oligonucleotide primers generated amplicons in presence of the test DNA solution indicating an inhibition-free PCR (Data not shown).

**Figure 1:**
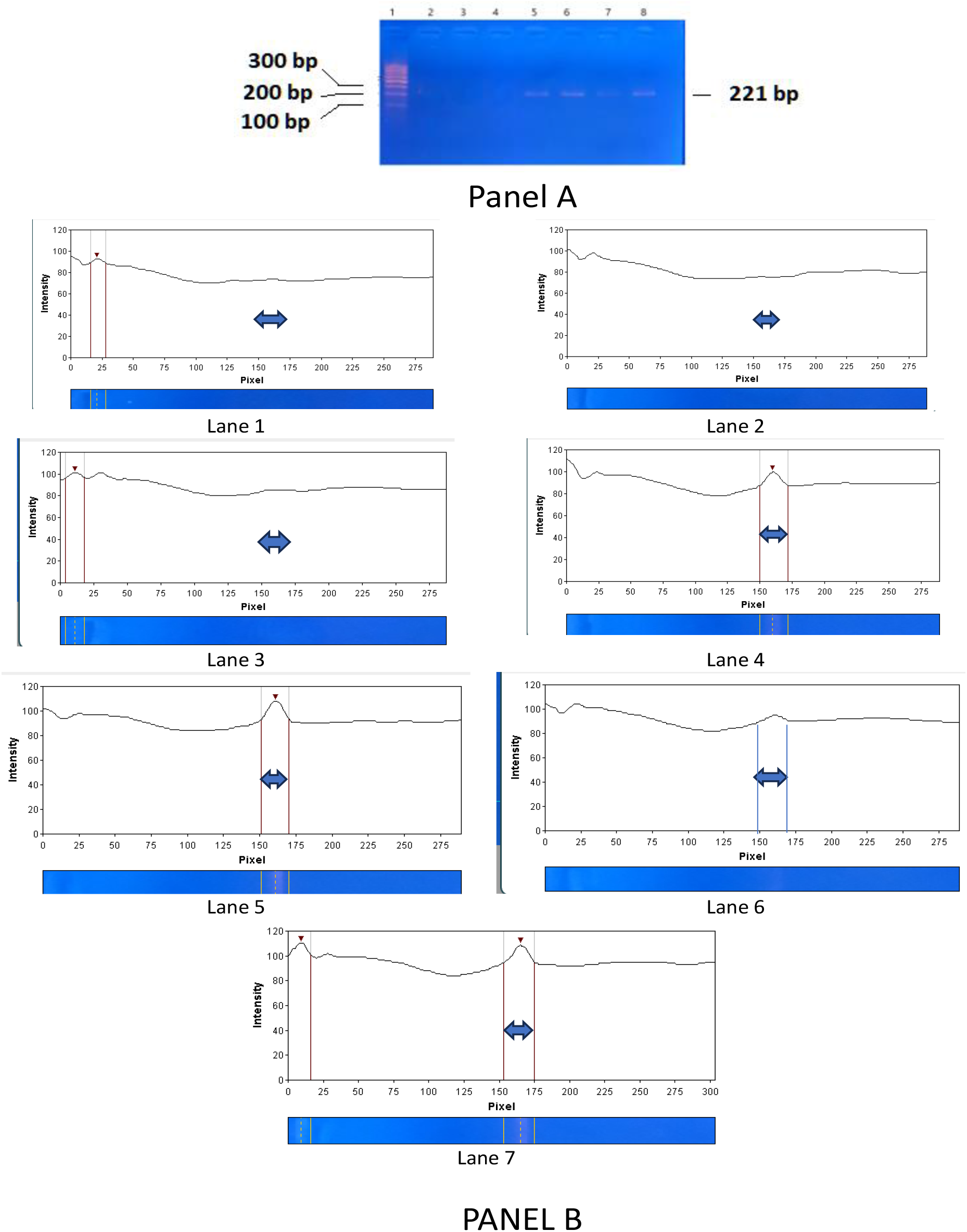
Agarose gel electrophoresis analysis of a panel of 7 M. tuberculosis-positive samples tested as ‘MTB Detected – Low’ by Xpert MTb Rif Ultra test. **Panel A:** Lane 1: 100 bp DNA size standard; 2-8: 5 μL PCR product from a 25 μL reaction volume resolved on the gel for samples XLw-1 to XLw-7 respectively. Sample XLw-1 to XLw-3 did not generate detectable band while CLw-4 – XLw-7 generated a faint band of size 221 bp. **Panel B:** Agarose gel lane scanning data using GelAnalyzer 23.1.1 software. Target bands were expected in the region between 150- and 175-Pixel points (X axis) that corresponded to 221 bp and shown in the graph using a double-headed arrow. No peaks were detected in Lane 1-3 that corresponded to samples XLw-1 - XLw-3 while small peaks were detected in Lane 4-7 that corresponded to samples XLw-4 - XLw-7 respectively.

The DNA from three end point PCR-negative samples were then processed for LPA as per prescribed protocol. Results indicated that out of them one was faintly LPA-positive (XLw-2) while the remaining two (XLw-1 and XLw-3) were LPA-negative (Figure 2). Five μL of the stock DNA from these two LPA-negative samples (XLw-1 and XLw-3) were subjected to whole genome amplification (WGA) and 5 μL of the whole genome amplified product was directly used for LPA. Five μL of the WGA product was also subjected to end point PCR (25 μL reaction volume) using ITS-specific primers and 5 μL of the PCR-amplified product was resolved on agarose gel electrophoresis. Intense bands of expected size (221 bp) were detected in both the samples and their band intensity was significantly high (Figure 3, Panel A & Panel B). Scanning of the lanes of agarose gel corresponding to the samples showed nil Pixel units for samples XLw-1 to XLw-3, 108, 110, 104, 110 for samples XLw-4 to XLw-7 and 160 and 174 for whole genome amplified XLw-1 and 3 samples respectively (Figure 3, Panel C). Samples XLw-1 and XLw-3 were also found to be LPA-positive post whole genome amplification and further, their drug sensitivity profile matched the original Xpert MTB Rif Ultra test profile of the samples (Figure 4).

**Figure 2:**
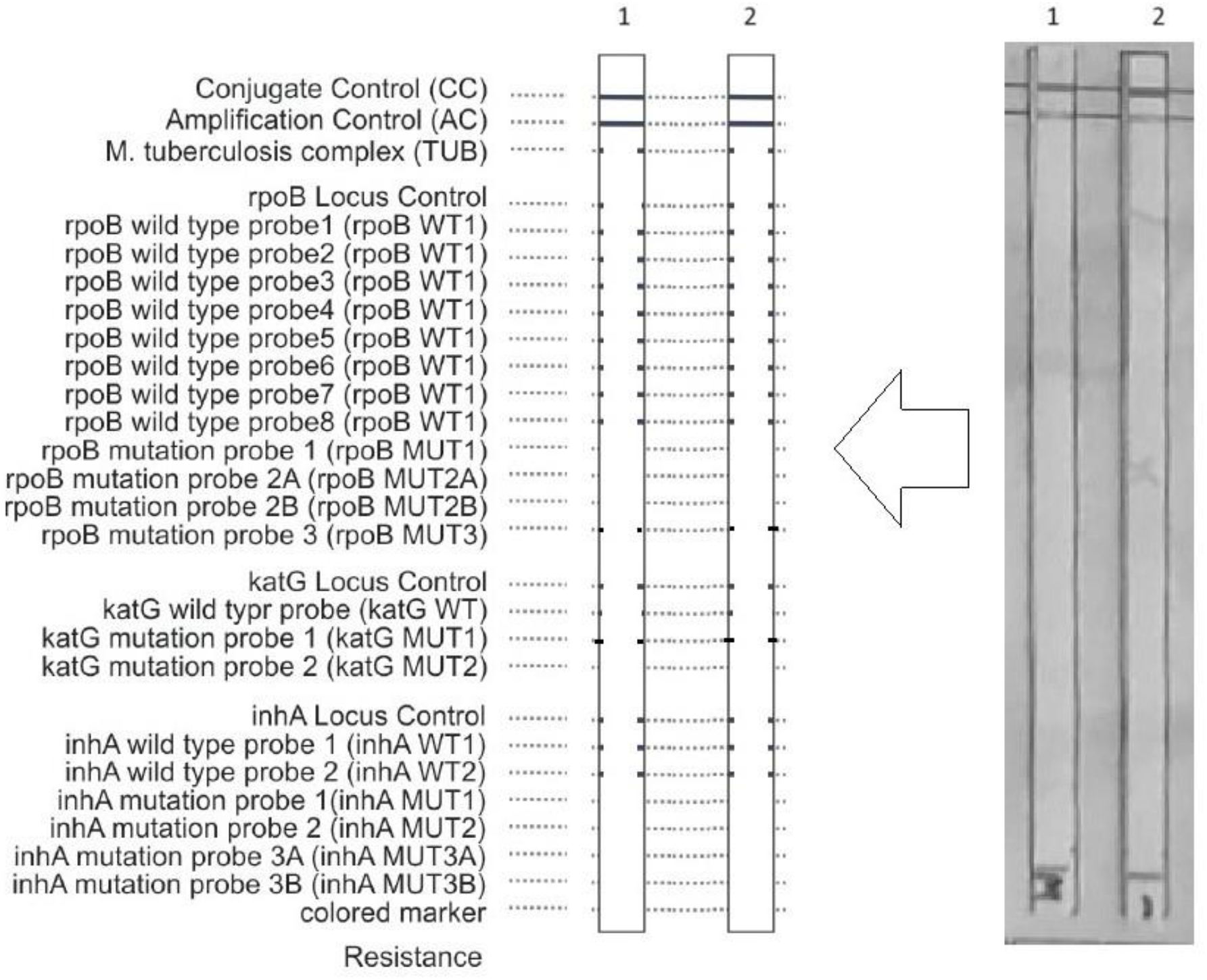
Line Probe Assay (LPA) for sample number XLw-1 (Strip 1) and XLw-3 (Strip 2) generated a *M. tuberculosis*-negative result due to low quantity of template DNA available for the assay. Only the conjugate and amplification controls are detected on top of the strips while all other signals are absent. The description in the extreme left indicates the interpretation of each band that is visible when optimal quantity of template DNA and the corresponding mutation is present or absent. The representative image in the middle is a sketch of the true result generated on the strip (extreme right).

**Figure 3:**
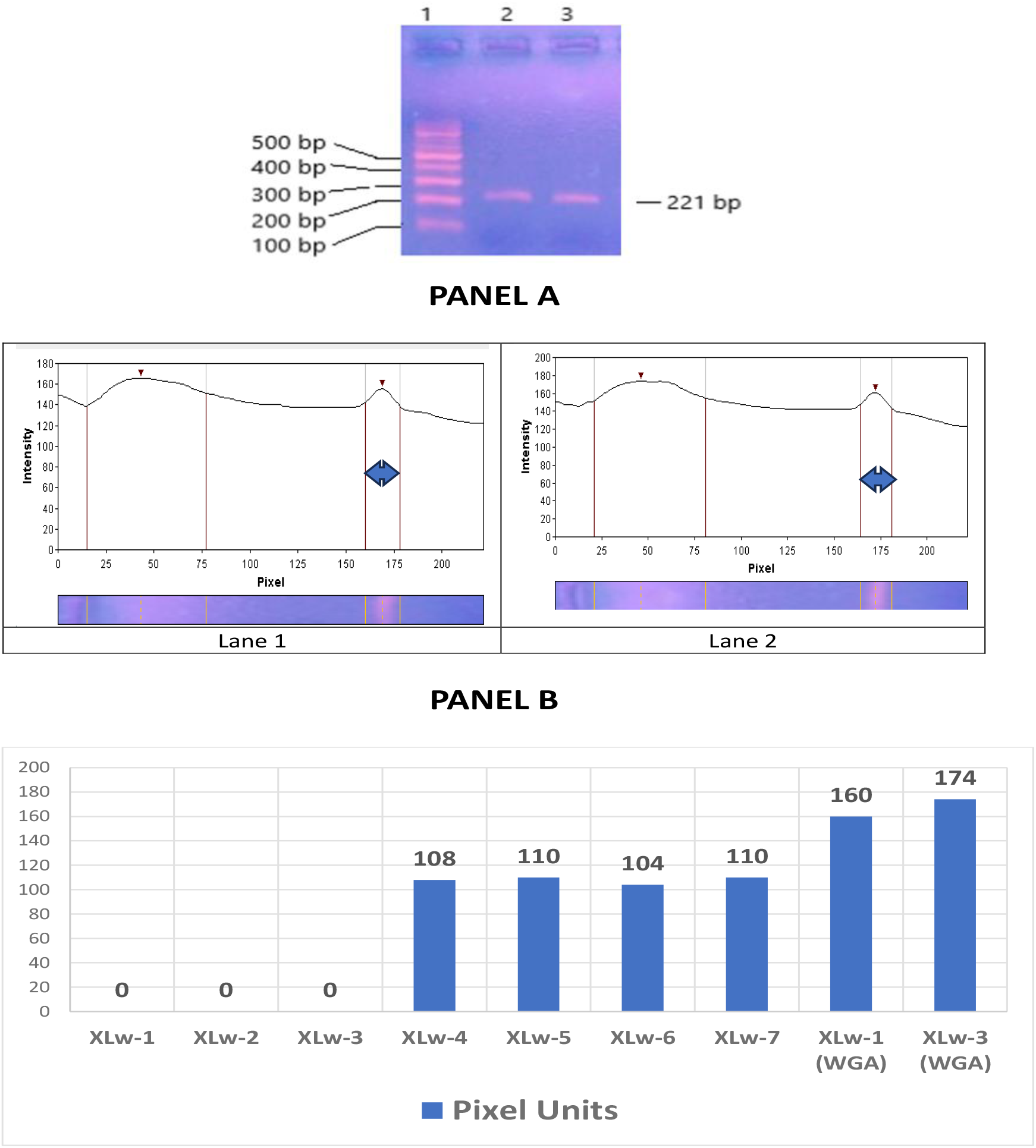
Agarose gel electrophoresis analysis of 5 μL of Whole Genome Amplified products (total reaction volume: 25 μL) resolved on agarose gel electrophoresis for sample no XLw-1 and XLw-3 respectively. **Panel A**: lane 1: 100 bp DNA size standard; Lane 2 & 3: Whole genome amplified products of samples XLw-1 and XLw-3 respectively. **Panel B:** Agarose gel (Panel A, this figure) lane scanning data using GelAnalyzer 23.1.1 software. Target bands were expected in the region between 162- and 177-Pixel points (X axis) that corresponded to 221 bp and shown in the graph using a double-headed arrow. Peaks were detected in Lane 1 & 2 that corresponded to whole genome amplified product of samples XLw-1 & XLw-3 respectively. **Panel C**: Graphical representation of the Fluorescent Intensity Units obtained from scanning of agarose gel electrophoresis lanes when end point PCR amplification products targeting the ITS (Internal Transcribed Spacer) region of M. tuberculosis genome were resolved. X axis: Sample XLw-1 to XLw-7 - Fluorescence obtained from agarose gel electrophoresis (Figure 1, Panel A); Sample XLw-1 (WGA) and XLw-3 (WGA) -Fluorescence obtained from agarose gel electrophoresis when whole genome amplified product of sample XLw-1 and 3, labelled as XLw-1 (WGA) and XLw-3 (WGA) were resolved. In all cases, 5 μL of the products were resolved from a 25 μL PCR or whole genome amplified reaction volume.

**Figure 4:**
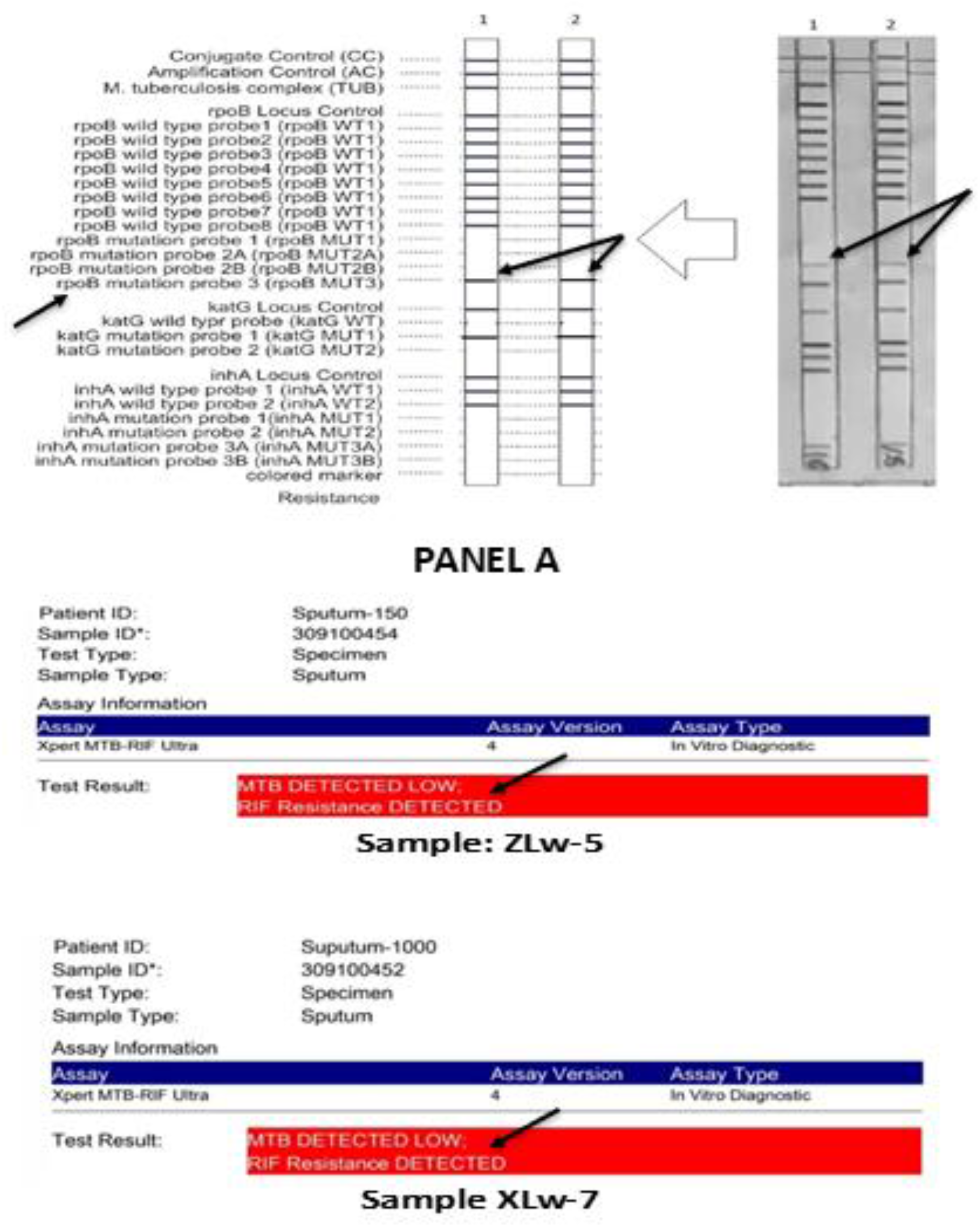
Results of Line Probe assay (LPA) of samples XLw-1 and XLw-3 after 5 μL of the original stock DNA released from cellulose matrix was subjected to Whole genome Amplification in a total reaction volume of 25 μL and 5 μL was used for running LPA. **Panel A:** Line Probe Assay (LPA) for Whole Genome Amplified version of sample number XLw-1 (Strip 1) and XLw-3 (Strip 2) which generated a *M. tuberculosis*-positive result with a drug resistant mutation corresponding to rpoB mutation probe 2B. The description in the extreme left indicates the interpretation of each band that is visible. The representative image in the middle is a sketch of the true result generated on the strip (extreme right). Panel B: Clip showing Xpert MTB Rif Ultra report of original stock of samples XLw-1 and XLw-3 respectively. The mutation profile observed in Panel A and Panel B for the samples XLw-1 and XLw-3 are identical (Panel A & B, Black solid arrows) the difference being that in Panel A, the data was obtained after Whole Genome Amplification of the stock DNA while in Panel B, the data was obtained from the original stock DNA.

The LPA reaction generated positive data when 5 μL of Dilution-1 samples (*M. tuberculosis* DNA of amount 125 ng spiked to *M. tuberculosis* -negative sputum DNA) were processed. However, when the same volume of dilution-2 DNAs (*M. tuberculosis* DNA of amount 1.25 ng spiked to *M. tuberculosis* negative sputum DNA) were processed, LPA returned a negative sample data. When 5 μL of dilution-2 DNA was subjected to whole genome amplification and same volume (5 μL) of the amplified products used to run LPA, the result was positive ((Figure 5) and the resistance/sensitivity profile of the samples was identical to that obtained from Dilution-1 sample (Figure 5).

**Figure 5:**
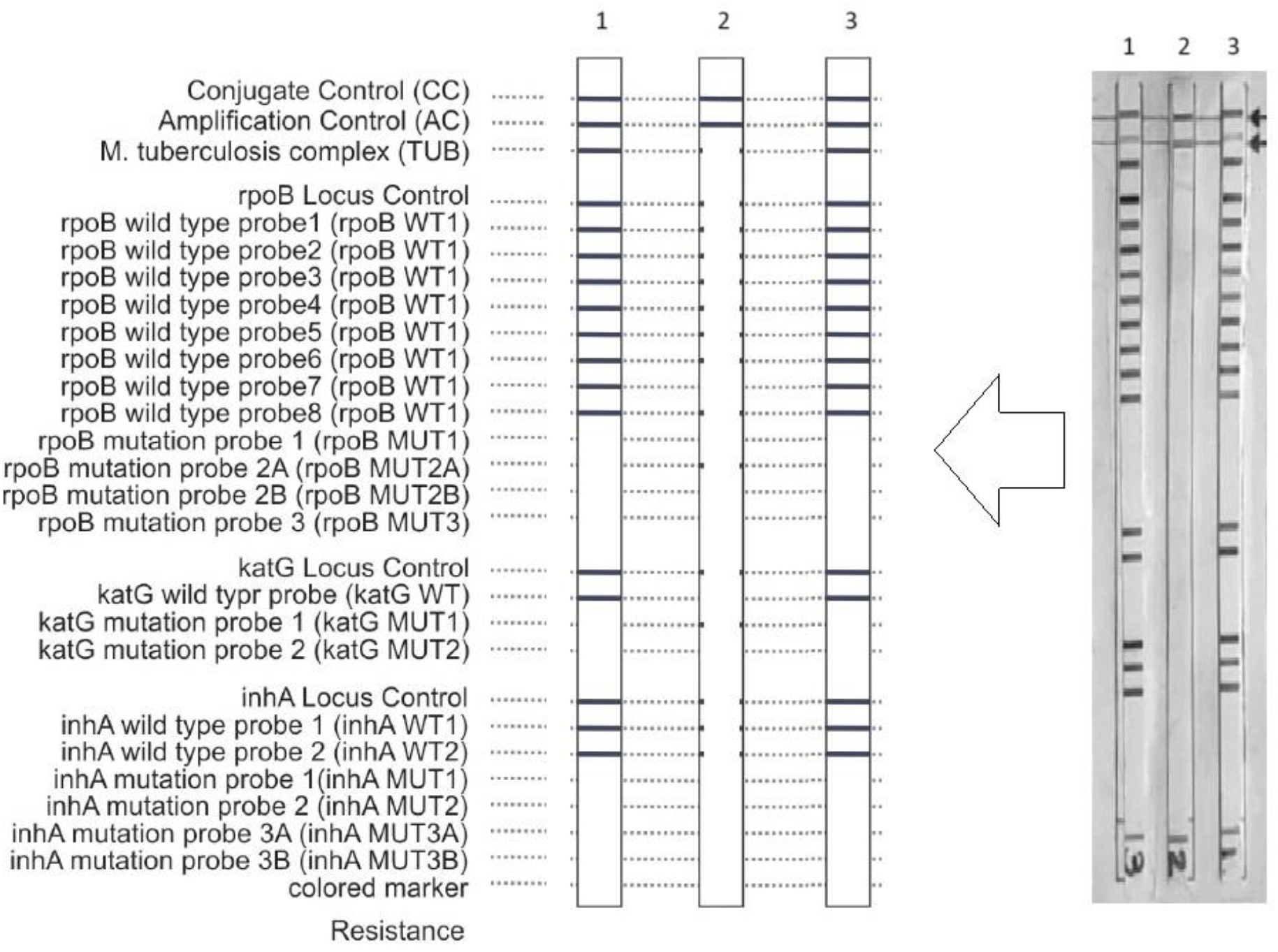
Results of Line Probe Assay (LPA) for Dilution 1 (Strip 1), Dilution 2 (Strip 2) and Whole genome Amplified Dilution 1 samples. Dilution 1 sample (125 ng *M. tuberculosis* DNA; Strip 1) and Whole genome Amplified Dilution 2 sample (Strip 3) generated identical *M. tuberculosis*-positive results while Dilution 2 (1.25 ng *M. tuberculosis* DNA; Strip 2) failed to generate *M. tuberculosis*-positive result. The description in the extreme left indicates the interpretation of each band that is visible when optimal quantity of template DNA and the corresponding mutation is present or absent. The representative image in the middle is a sketch of the true result generated on the strip (extreme right). The dilution samples were formulated by adding precise quantity of DNA extracted from a laboratory culture of *M. tuberculosis* strain to DNA extracted from sputum of healthy individuals.

## Discussion

Simultaneous testing of isoniazid and rifampicin resistance when diagnosis of tuberculosis is performed is perhaps the most economical approach at all settings to increase the rate of cure, reduce mortality and stunt the emergence of drug resistance in cases of tuberculosis by reducing the chances of failure and relapse [16]. In the past few decades tuberculosis diagnosis saw a mammoth revolution in molecular diagnostic techniques to detect sensitive and resistant variants of *M. tuberculosis*. The threatening emergence of drug resistance in tuberculosis pose a strong challenge to effective delivery of healthcare solutions at a global level and call from not only development of new antimicrobials but also detection of drug resistant variants of the pathogen in a way that is easily accessible, economical and sensitive [17].

The line probe assay or LPA is a very promising solution for rapid detection of *M. tuberculosis* drug resistance. It exploits the principle of multiplex polymerase chain reaction and reverse hybridization assay [18]. The World Health Organization (WHO) has endorsed LPA for adoption in screening and diagnostic algorithms for pulmonary tuberculosis in countries with high burden of the disease [18].

Collection of clinical samples for tuberculosis diagnosis continues to remain an under-focussed aspect in the entire ecosystem of molecular diagnosis of the disease. Numerous challenges are associated with conventional methods of collection, transportation and storage of sputum samples prior to molecular diagnosis [19]. These range from leakage of infectious samples to extensive contamination while on long transit. In this scenario, a solid matrix for room temperature transportation as well as storage, coupled with easy release of DNA present within the heterogeneous cellular population of the clinical sample trapped onto it, positions itself as a useful and viable alternative [11].

It is therefore important to demonstrate seamless integration of such solid-support DNA storage devices with detection technologies such as LPA such that tuberculosis diagnosis gains further strength to cover larger geographical areas of countries with high burden of the disease such as India. This study uses such a sputum transportation, storage and DNA release device [11], [12] and demonstrate its integration with LPA *via* a short but effective whole genome amplification step that significantly enhances the sensitivity of LPA.

Hain Lifescience (GmbH, Nehren, Germany), manufacturer of a popular LPA product for detecting drug resistant tuberculosis recommends a volume of 5 μL of DNA with concentration ranging between 20-50 ng/μL as the starting material for running the assay. Therefore, a total amount of 100-250 ng of DNA is recommended for proper functioning of the assay. However, it is not always feasible to get a *M. tuberculosis* positive sample of desired optimal concentration for LPA which can increase the probability of generating false negatives on this platform.

In order to address this issue, an approach was adopted wherein sub optimal quantity of DNA, (with regards to LPA assay) released from a solid support as described by Goyani et al., (2023) [11], [12], was amplified by use of *Bst* DNA polymerase wherein its strong strand displacement activity was exploited to achieve long and continuous stretch of amplified DNA in a short period of time.

While other popular methods of whole genome amplification, such as Multiple Displacement Amplification (MDA) using phi29 DNA polymerase [20] and Degenerate Oligonucleotide-Primed PCR (DOP-PCR) employing degenerate primers that prime randomly across the genome [21] are available and are used more frequently in the molecular biotechnology field, the current method proved to be effective in this particular case owing to the small size of the target-genome compared to that in human cells, that got preferentially amplified during the process.

The experiment demanded clinical sputum samples with low bacterial pathogen load. Earlier studies indicate that bacterial load of *M. tuberculosis* in sputum sample of patients who are under treatment is low which corresponds with improved clinical conditions and reduced transmission risk of transmission [22]. This was the rationale behind specific composition of the resource population used in this study where all samples obtained were from tuberculosis-patients undergoing 1^st^ line of therapy (Isoniazid, Rifampicin, Pyrazinamide, and Ethambutol or HRZE) [23].

Screening of 39 patients yielded 7 samples that recorded “MTB detected low” when tested on Xpert MTB Rif Ultra assay. This invariably indicated low load of pathogen in the clinical sample and hence low pathogen DNA in the corresponding extracted DNA samples. This observation was reflected when the extracted DNA from these 7 samples (XLw-1 to XLw-7) were subjected to a traditional end point PCR targeting the ITS region of the pathogen genome. As expected, 3 samples (XLw-1 to XLw-3) failed to generate any detectable PCR amplicons when resolved on an agarose gel while remaining 4 samples (XLw-4 to XLw-7) generated faint bands (Figure 1). This result was expected since previous studies show that *M. tuberculosis* DNA in sputum samples that generate low signals in the Xpert MTB Rif Ultra assay (or its variants) are most often not detectable by end point PCR methods [24].

These 3 samples, *viz*., XLw-1, 2 and 3 respectively, were taken for further study and subjected to Line Probe Assay. Our data show that while the LPA test was weakly positive for sample XLw-2, samples XLw-1 and XLw-3 were reported as false negatives (Figure 2). This observation is in line with earlier findings that indicate that sensitivity of line probe assays for detecting *M. tuberculosis* genome targets is less when compared to fluorescent real - time PCR methods [25].

When the stock DNA of samples XLw-1 and XLw-3 were subjected to whole genome amplification followed by end point PCR targeting the ITS region of the *M. tuberculosis* genome, intense bands were detected (Figure 3). It may be noted that in all agarose gel electrophoresis experiments, a total volume of 5 μL of DNA was loaded after adding 2 μL of agarose gel loading dye (6X). Given the fact that 5 μL of stock DNAs of Samples XLw-1 and XLw-3 were used for whole genome amplification in a total volume of 25 μL and 5 μL of the whole genome amplified product was resolved in the agarose gel, it effectively corresponded to 1/5th or 1 μL of the stock DNA. Relative fluorescence of the PCR amplicons after agarose gel electrophoresis indicate that the intensity of the whole genome amplified products (post-WGA, DNA-amplified products; sample XLw-1 WGA and XLw-3 WGA) were 160- and 174-Pixel units as compared to XLw-4, 5, 6 and 7 samples (pre-WGA stage, stock DNA-amplified products) where the band intensity ranged between 104 and 110 Pixel units and nil in case of samples XLw-1, 2 and 3 (pre-WGA stage, stock DNA-amplified products) (Panel C, Figure 3).

This showed that the genome region targeted by the conventional end point PCR got sufficiently amplified during the whole genome amplification process and provided the desired quantity of template DNA for successful PCR amplification in samples XLw-1 (WGA) and XLw-3 (WGA).

Whole genome amplification (WGA) employing the Bst DNA polymerase occurs through Multiple Displacement Amplification (MDA) which is known to be a highly efficient process for amplifying small quantities of DNA into microgram levels with high degree of fidelity. This method has earlier been demonstrated for whole genome amplification in other biological samples [26].

Post agarose gel electrophoresis analysis, the whole genome amplified products corresponding to samples XLw-1 and XLw-3 were subjected to LPA. Results indicated that both the amplified samples effectively generated positive LPA results and further, the drug resistance profile matched with that obtained from Xpert MTB Rif Ultra tests performed on the original stock DNA of these two samples (Figure 4).

In order to confirm the effectiveness of the whole genome amplification step, DNA extracted from sputum of healthy individuals were spiked with *M. tuberculosis* DNA extracted from *M. tuberculosis* bacilli cultured in the laboratory in LJ medium [27]. DNA extracted from human sputum samples contains both mycobacterial as well as human DNA which can pose complications in molecular diagnostic processes during downstream analysis [28]. Therefore, in order to mimic a typical infected sputum sample, this *M tuberculosis* DNA-spiked healthy sputum DNA was formulated.

Results indicated that in sample coded Dilution-1 where the total *M. tuberculosis* DNA available for LPA reaction was 125 ng, a positive LPA result was obtained. However, when the sample coded Dilution-2 containing 1.25 ng of total *M. tuberculosis* DNA was subjected to LPA, a false negative report was generated.

When 5 μL of the Dilution-2 DNA was subjected to 10 minutes of whole genome amplification process and 5 μL of the amplified product used to run an LPA, the result was positive and further, the drug resistance /sensitivity profile matched that of Dilution-1 sample. This indicated that the process of amplification of the specific targets within the whole genome of *M. tuberculosis* present in the Dilution-2 formulation occurred effectively and efficiently to return a positive LPA result.

Based on the findings presented in this paper, it is evident that addressing the challenge of low sensitivity in Line Probe Assay (LPA) compared to fluorescent real-time PCR methods can be effectively achieved through the innovative use of sputum DNA archived in chemically coated solid, cellulose matrixes. The matrix facilitates room temperature transportation and extended storage of clinical samples, while its ultra-short DNA release step and brief whole genome amplification protocol efficiently amplify low quantities of *M. tuberculosis* DNA present in these samples. The application of this technology not only enhances the sensitivity of *M. tuberculosis* LPA but also has the potential to convert false negative results into true positive outcomes. This approach represents a significant advancement in diagnostic capabilities, particularly in resource-limited settings where maintaining cold chain logistics is challenging. By improving the detection limit and reliability of LPA, the integration of such sputum transportation, storage and DNA-release technology promises to contribute positively to the diagnosis and management of tuberculosis.

## Conclusion

In conclusion, leveraging the cellulose matrix technology for room temperature transportation, extended storage, and enhanced DNA release and its integration with a brief whole genome amplification protocol offers a promising solution to enhance the sensitivity of *M. tuberculosis* LPA, thereby improving the accuracy and effectiveness of tuberculosis diagnostics.

## Data Availability

All data produced in the present work are contained in the manuscript

## Acknowledgements

This study was partly supported by Grand Challenge-TB Control (GCTBC) grants, namely GCTBC/C2P1/2015/09/30/01 & GCTBC/C2P2/2017/04/01/01 and co-funded by Industry Research Assistance Council (BIRAC) of the Government of India, United States Agency for International Development (USAID), and Bill & Melinda Gates Foundation. IKP Knowledge Park (Hyderabad, India) was the implementation partner for these programmes.

## References

1. World Health Organization. Global Tuberculosis Report. Geneva, World Health Organization, 2016. Available from: http://www.who.int/tb/publications/global_report/en/

2. World Health Organization. Multidrug and Extensively Drug-resistant TB (M/XDR-TB): 2010 Global Report on Surveillance and Response. Geneva, World Health Organization, 2010. Available from: http://apps.who.int/iris/bitstream/10665/44286/1/9789241599191_eng.pdf

3. Heifets, L. B., & Cangelosi, G. A. (1999). Drug susceptibility testing of Mycobacterium tuberculosis: a neglected problem at the turn of the century. The international journal of tuberculosis and lung disease: the official journal of the International Union against Tuberculosis and Lung Disease, 3(7), 564–581.

4. Dinnes, J., Deeks, J., Kunst, H., Gibson, A., Cummins, E., Waugh, N., Drobniewski, F., & Lalvani, A. (2007). A systematic review of rapid diagnostic tests for the detection of tuberculosis infection. Health technology assessment (Winchester, England), 11(3), 1–196.

5. Small, P. M., & Pai, M. (2010). Tuberculosis diagnosis--time for a game change. The New England journal of medicine, 363(11), 1070–1071.

6. Genotype MTBDRplusTM, version 1.0 [product insert]. Nehren, Germany: Hain Life science, GmbH. Hain life science website. Available: http://www.hainlifescience.com/pdf/304xx_pbl.pdf. Accessed 2013 Nov 1

7. Leary, J. J., Brigati, D. J., & Ward, D. C. (1983). Rapid and sensitive colorimetric method for visualizing biotin-labeled DNA probes hybridized to DNA or RNA immobilized on nitrocellulose: Bio-blots. Proceedings of the National Academy of Sciences of the United States of America, 80(13), 4045–4049.

8. Roscigno, G. (2009). Drug resistant TB and new diagnostics for people living with HIV: emerging results from FIND. IAS Conference; Catalysing HIV/TB research.

9. Lawn, S. D., & Nicol, M. P. (2011). Xpert® MTB/RIF assay: development, evaluation and implementation of a new rapid molecular diagnostic for tuberculosis and rifampicin resistance. Future microbiology, 6(9), 1067–1082.

10. Ninan, M. M., Gowri, M., Christopher, D. J., Rupali, P., & Michael, J. S. (2016). The diagnostic utility of line probe assays for multidrug-resistant tuberculosis. Pathogens and global health, 110(4-5), 194–199.

11. Goyani, K. H., & Mukhopadhyaya, P. N. (2023). Sputum-Spotted Solid Matrix Designed to Release Diagnostic-Grade Mycobacterium tuberculosis DNA Demonstrate Optimal Biocontainment Property. International Journal of Science and Research (IJSR), 12(10), 1082–1088.

12. Goyani, K. H., Mukhopadhyaya, P. N. (2023). Estimating Limit of Detection of M. Tuberculosis DNA as an Analyte Released from a Chemically Coated Solid Matrix and Using a WHO-approved CB-NAAT Platform. Archives of Infect Diseases & Therapy, 7(3), 79–84.

13. Chakravorty, S., Simmons, A. M., Rowneki, M., Parmar, H., Cao, Y., Ryan, J., Banada, P. P., Deshpande, S., Shenai, S., Gall, A., Glass, J., Krieswirth, B., Schumacher, S. G., Nabeta, P., Tukvadze, N., Rodrigues, C., Skrahina, A., Tagliani, E., Cirillo, D. M., Davidow, A., … Alland, D. (2017). The New Xpert MTB/RIF Ultra: Improving Detection of Mycobacterium tuberculosis and Resistance to Rifampin in an Assay Suitable for Point-of-Care Testing. mBio, 8(4), e00812–17.

14. Lasken, R. S., & Egholm, M. (2003). Whole genome amplification: abundant supplies of DNA from precious samples or clinical specimens. Trends in biotechnology, 21(3), 153–158.

15. Middlebrook, G., & Cohn, M. L. (1958). Bacteriology of tuberculosis: laboratory methods. American journal of public health and the nation’s health, 48(7), 844–853.

16. WHO Guidelines for the programmatic management of drug resistant tuberculosis, 2011. [Internet]. [cited 2011 Nov 8]. Available from: http://www.who.int/tb/challenges/mdr/programmatic_guidelines_for_mdrtb/en/

17. Nathan C. (2020). Resisting antimicrobial resistance. Nature reviews. Microbiology, 18(5), 259–260.

18. World Health Organisation: Policy statement. Molecular Line Probe Assays for Rapid Screening of patients at risk of multidrug resistant tuberculosis (MDR-TB); 2008.

19. Wares, D. F., Akhtar, M., Singh, S., & Jain, N. K. (2001). Bringing tuberculosis services to the underserved: a field report from rural India. International Journal of Tuberculosis and Lung Disease, 5(4), 336–344.

20. Dean, F. B., Hosono, S., Fang, L., Wu, X., Faruqi, A. F., Bray-Ward, P., Sun, Z., Zong, Q., Du, Y., Du, J., Driscoll, M., Song, W., Kingsmore, S. F., Egholm, M., & Lasken, R. S. (2002). Comprehensive human genome amplification using multiple displacement amplification. Proceedings of the National Academy of Sciences of the United States of America, 99(8), 5261–5266.

21. Telenius, H., Carter, N. P., Bebb, C. E., Nordenskjöld, M., Ponder, B. A., & Tunnacliffe, A. (1992). Degenerate oligonucleotide-primed PCR: general amplification of target DNA by a single degenerate primer. Genomics, 13(3), 718–725.

22. Samb, B., Sow, P. S., Kony, S., Maynart, M., Bakker, M. I., Sylla, O., … & Colebunders, R. (1999). Mycobacterium tuberculosis load in sputum of patients with pulmonary tuberculosis undergoing treatment: Correlation with clinical improvement. Journal of Clinical Microbiology, 37(12), 3834–3836.

23. Sachdeva, K. S., Raizada, N., Sreenivas, A., Van’t Hoog, A. H., Nair, S. A., Kulsange, S., … & Paramasivan, C. N. (2012). Use of Xpert MTB/RIF in Decentralized Public Health Settings and Its Effect on Pulmonary TB and DR-TB Case Finding in India. PLOS ONE, 7(5), e37631.

24. Theron, G., Peter, J., van Zyl-Smit, R., Mishra, H., Streicher, E., Murray, S., … & Dheda, K. (2011). Evaluation of the Xpert MTB/RIF assay for the diagnosis of pulmonary tuberculosis in a high HIV prevalence setting. American Journal of Respiratory and Critical Care Medicine, 184(1), 132–140.

25. Lawn, S. D., Mwaba, P., Bates, M., Piatek, A., Alexander, H., Marais, B. J., & Bates, M. N. (2013). Advances in tuberculosis diagnostics: The Xpert MTB/RIF assay and future prospects for a point-of-care test. The Lancet Infectious Diseases, 13(4), 349–361.

26. Dean, F. B., Hosono, S., Fang, L., Wu, X., Faruqi, A. F., Bray-Ward, P., … & Lasken, R. S. (2002). Comprehensive human genome amplification using multiple displacement amplification. Proceedings of the National Academy of Sciences, 99(8), 5261–5266.

27. Kent & Kubica. (1985). Lowenstein-Jensen (LJ) medium is widely used for the cultivation and detection of Mycobacterium tuberculosis due to its efficacy in supporting the growth of mycobacteria while inhibiting contaminants.

28. Hance, A. J., Grandchamp, B., Levy-Frebault, V., Lecossier, D., Rauzier, J., Bocart, D., & Gicquel, B. (1989). Detection and identification of mycobacteria by amplification of mycobacterial DNA. Molecular Microbiology, 3(7), 843–849.

